# Explainable Deep Learning for Lesion-Level Detection of Diabetic Retinopathy: A Segmentation Approach Using Fundus Images Graded as Mild-to-Moderate Nonproliferative Diabetic Retinopathy

**DOI:** 10.1101/2025.10.01.25337115

**Authors:** Takumi Sato, Koichi Nishitsuka, Tohru Itoh, Toshihiro Okashita, Satoshi Wada, Atsushi Shinjo

## Abstract

Deep learning has shown promise in diabetic retinopathy screening using fundus images. However, many existing models operate as “black boxes,” providing limited interpretability at the lesion level. This study aimed to develop an explainable deep learning model capable of detecting four diabetic retinopathy-related lesions—hemorrhages, hard exudates, cotton wool spots, and microaneurysms—and evaluate its performance using both conventional per-lesion metrics and a novel syntactic agreement framework. A total of 1,087 fundus images were obtained from publicly available datasets (EyePACS and APTOS), which contained 585 images graded as mild-to-moderate nonproliferative diabetic retinopathy (DR1 or DR2). All images were manually annotated for the presence of the four lesions. A U-Net–based segmentation model was trained to generate binary predictions for each lesion type. The performance of the model was evaluated using sensitivity, specificity, precision, and F1 score, along with five syntactic agreement criteria that evaluated the lesion-set consistency between the predicted and ground truth outputs at the image level. The model achieved high sensitivity and F1 scores for hemorrhages and hard exudates, showed moderate performance for cotton wool spots, and failed to detect any microaneurysms (0% sensitivity), with 92.9% of the microaneurysms cases misclassified as hemorrhages. Despite this limitation, the image-level agreement remained high, with any-lesion match and hemorrhage match rates exceeding 95%. These findings suggest that although individual lesion classification was imperfect, the model effectively recognized abnormal images, highlighting its potential as a screening tool. The proposed syntactic agreement framework offers a complementary evaluation strategy that aligns more closely with clinical interpretation and may help bridge the gap between artificial intelligence–based predictions and real-world ophthalmic decision-making.

## Introduction

Diabetic retinopathy is a leading cause of vision loss globally, particularly among working-age adults [1]. Early detection and timely treatment are crucial for preventing irreversible visual impairment. However, with the increasing global burden of diabetes and the shortage of ophthalmologists in many regions, deep learning-based automated retinal image analysis has emerged as a promising strategy for scalable and efficient diabetic retinopathy screening [2,3].

Recent advances in deep learning have demonstrated that convolutional neural networks can detect diabetic retinopathy from fundus photographs with high accuracy, with the accuracy sometimes matching or even surpassing that of human experts [2,3]. However, most of these models operate as “black boxes,” outputting only an overall diabetic retinopathy severity grade without identifying the specific retinal lesions that drive the output. This lack of interpretability hinders clinical trust and poses a barrier to the integration of artificial intelligence (AI) tools into real-world ophthalmic practice [4].

To improve model transparency, lesion-level detection approaches have been developed to localize specific pathological features, such as hemorrhages, hard exudates, and microaneurysms [6,7]. These methods enhance interpretability and help clinicians better understand the rationale behind AI-driven outputs. However, their evaluation typically relies on pixel-level per-lesion performance metrics—such as sensitivity, precision, and F1 score—which assess each lesion type in isolation. Although these metrics are useful, they may fail to capture the overall clinical relevance of the model’s image-level prediction.

In clinical practice, ophthalmologists interpret fundus images holistically—considering not only the pixel-level morphology of individual lesions but also their combinations and spatial distribution—to assess disease severity. To better emulate this diagnostic reasoning, we propose a syntactic agreement framework that evaluates the consistency between the predicted and reference lesion sets at the image level. The proposed framework offers a complementary perspective to traditional per-lesion metrics and enables a more clinically meaningful evaluation of AI-based predictions.

In this study, we developed a deep learning model for the lesion-level detection of diabetic retinopathy using publicly available fundus images. We evaluated the performance of the proposed model using both per-lesion metrics and a novel syntactic agreement framework to assess image-level consistency across the diabetic retinopathy stages. Additionally, we conducted qualitative analyses of model misclassifications to explore common failure patterns and highlight challenges in lesion-specific interpretability.

## Methods

### Dataset and Image Selection

We used a total of 1,087 de-identified color fundus photographs obtained from the publicly available datasets EyePACS and APTOS, which are widely used in diabetic retinopathy-related AI research. The EyePACS dataset was accessed on June 20, 2024, and the APTOS dataset was accessed on June 20, 2024, for research purposes. Authors did not have access to information that could identify individual participants during or after data collection. The datasets used (EyePACS and APTOS) were fully anonymized and provided without any personally identifiable information. All images were captured using nonmydriatic fundus cameras (Topcon Corporation, Tokyo, Japan) to ensure consistency in image resolution and quality. For primary analysis, we selected 585 images from patients diagnosed as mild-to- moderate nonproliferative diabetic retinopathy, corresponding to DR1 and DR2 stages according to the International Clinical Diabetic Retinopathy Disease Severity Scale [5]. In this scale, DR0 indicates no apparent retinopathy; therefore, DR0 images were excluded from the main analysis. Images from patients diagnosed as early-stage diabetic retinopathy (DR1 and DR2) were selected because they pose significant challenges for conventional deep learning classifiers [2]. Each selected image was independently reviewed and annotated for the presence of four key diabetic retinopathy-related lesions: hemorrhages, hard exudates, cotton wool spots, and microaneurysms.

### Lesion Annotation and Ground Truth

Lesion annotations were conducted using a custom-designed image segmentation tool. The annotator was a researcher with substantial experience in fundus photography, working under the supervision of a board-certified ophthalmologist specializing in retinal image interpretation. Each lesion was labeled at the pixel level using binary masks for four predefined categories: hemorrhages, hard exudates, cotton wool spots, and microaneurysms. The pixel-wise annotations were then converted into binary presence/absence labels for each lesion type on a per-image basis. These labels were used as the ground truth for the training and evaluation of the AI model. To ensure optimal data quality, images with poor visibility or ambiguous findings were excluded from the analysis. Representative examples of the annotated masks are presented in Supplementary Figure S1.

### Image Preprocessing

All fundus images were resized to 512 × 512 pixels and rescaled to the [0, 1] intensity range. Channel-wise normalization was then applied using ImageNet statistics (mean of RGB: [0.485, 0.456, 0.406]; standard deviation of RGB: [0.229, 0.224, 0.225]), consistent with the pretrained encoder used in this study. To preserve anatomical laterality—such as the location of the optic disc—no horizontal flipping was applied during preprocessing. However, horizontal and vertical flipping were employed during model training as part of the data augmentation strategy to improve the generalization performance.

### Model Architecture and Training

We implemented a U-Net–based convolutional neural network for the multilabel segmentation of diabetic retinopathy lesions. The input comprised 512 × 512 RGB fundus images, and the model generated four binary segmentation masks, corresponding to hemorrhages, hard exudates, cotton wool spots, and microaneurysms. A sigmoid activation function was applied to the final layer to enable the independent binary classification of each lesion type.

The model was implemented in PyTorch and trained on NVIDIA GPUs using the Adam optimizer. The loss function was a Dice loss for the localization performance. Data augmentation techniques included horizontal and vertical flipping, brightness and contrast adjustment, and random rotation to enhance generalizability.

### Performance Evaluation

For each lesion type, the binary predictions (presence or absence) of the model were compared with the ground truth annotations to calculate the sensitivity, specificity, precision, and F1 score. These metrics were computed based on the number of true positives, false positives, false negatives, and true negatives evaluated on a per-image basis. In addition to quantitative assessment, we qualitatively analyzed cross-lesion misclassifications, including cases where microaneurysms were misclassified as hemorrhages, to understand the common error patterns. To avoid artificially inflated specificity values due to the large number of true negatives in DR0 images, all quantitative performance evaluations were restricted to images labeled as DR1 or DR2.

### Syntactic Agreement Evaluation

To complement the traditional lesion-wise performance metrics, we introduced five syntactic agreement criteria to evaluate the consistency between the AI-predicted and ground truth lesion sets at the image level. These criteria aim to approximate the clinical reasoning process by capturing the presence of individual lesions and their co-occurrence patterns. The syntactic agreement metrics were defined as follows:

- Strict match: The predicted lesion set exactly matches the ground truth set.
- Syntactic containment: The predicted lesion set is a nonempty subset of the ground truth sets, indicating conservative predictions without false positives. Images with no predicted lesions were marked as “Not Applicable.”
- Hemorrhage match: Assessed only in images with ground truth hemorrhages. A match is recorded if the AI prediction also includes the presence of hemorrhages. Images lacking hemorrhages in the ground truth were marked as “Not Applicable.”
- Lesion match: At least one predicted lesion overlaps with the ground truth. Applied only to images with one or more ground truth lesions (DR1 or DR2); images with no overlap were marked as mismatches.
- Any-lesion match: This evaluates whether the AI detected any lesion—regardless of correctness—in images with at least one annotated lesion. This metric assesses the model’s ability to flag an image as abnormal even when the lesion classification is imperfect.

For DR0 images (no annotated lesions), only “strict match” was applied to evaluate whether the AI correctly predicted no lesion outputs. All other syntactic criteria were deemed not applicable and were excluded from the evaluation. Table 1 presents the representative scenarios of how each criterion is applied. To illustrate how these criteria are applied across all lesion combinations, we generated 16 × 16 binary matrices by comparing the predicted and ground truth lesion codes. Each matrix visualizes whether a given syntactic criterion was satisfied (red), not satisfied (blue), or not applicable (blank). These comprehensive visualizations (Supplementary Figure S2A–E) offer intuitive insights into the relative stringency and overlap of the evaluation criteria.

**Table 1.** Representative scenarios for syntactic agreement evaluation using defined lesion-level criteria.

| Ground truth | AI output | Strict match | Syntactic containment | Hemorrhage match | Lesion match | Any-lesion match |
| --- | --- | --- | --- | --- | --- | --- |
| H, HE, CWS, MA | H, HE, CWS, MA | Yes | Yes | Yes | Yes | Yes |
| H, HE, CWS, MA | H, HE, CWS | No | Yes | Yes | Yes | Yes |
| H, CWS, MA | H, HE | No | No | Yes | Yes | Yes |
| H, CWS, MA | HE, CWS | No | No | No | Yes | Yes |
| H, HE, CWS, MA | None | No | No | No | No | No |
| MA | H | No | No | No | No | Yes |

This table demonstrates how five syntactic agreement criteria can be applied to various combinations of ground truth (GT) and artificial intelligence (AI)–predicted lesion sets. The criteria include strict match (exact correspondence between GT and AI), syntactic containment (AI predictions as a nonempty subset of GT), hemorrhage match (both GT and AI include hemorrhages), lesion match (at least one lesion type is shared), and any-lesion match (AI predicts any lesion when GT includes at least one). These examples illustrate the logical relationships and distinct thresholds among the criteria, facilitating the interpretation of lesion-wise evaluation strategies.
Abbreviations: H, hemorrhages; HE, hard exudates; CWS, cotton wool spots; MA, microaneurysms; GT, ground truth; AI, artificial intelligence.

## Results

### Lesion Prevalence and Ground Truth Distribution

Lesion-level analysis was performed on a subset of 585 fundus images from patients diagnosed with mild-to-moderate diabetic retinopathy (DR1 or DR2). Gound truth annotations were available for four key lesion types, with the following prevalence rates: hemorrhages (89.9%), hard exudates (41.2%), cotton wool spots (29.1%), and microaneurysms (12.0%). These distributions revealed a substantial imbalance across lesion categories, particularly highlighting the rarity of microaneurysms and cotton wool spots in the dataset.

### Lesion-wise Detection Performance

The model’s lesion-wise performance was assessed by comparing its binary predictions with the ground truth annotations for each lesion type. As shown in Table 2, the model achieved high sensitivity and F1 scores for hemorrhages and hard exudates while maintaining consistently high specificity across all lesion categories. In contrast, the detection performance was substantially poor for cotton wool spots and critically poor for microaneurysms, which were not correctly identified in any of the 70 ground truth- positive cases. These results highlight a significant limitation in the model’s ability to recognize subtle or less frequent lesions. Supplementary Table S1 provides a stratified analysis by diabetic retinopathy stage (DR1 vs. DR2), and Figure 2 presents a visual summary of the lesion-wise metrics. Figure 3 shows representative examples of the correctly detected lesions. Typical detection outcomes, including both accurate and erroneous cases, are further illustrated in Figure 4 and described in detail below.

**Fig 1.**
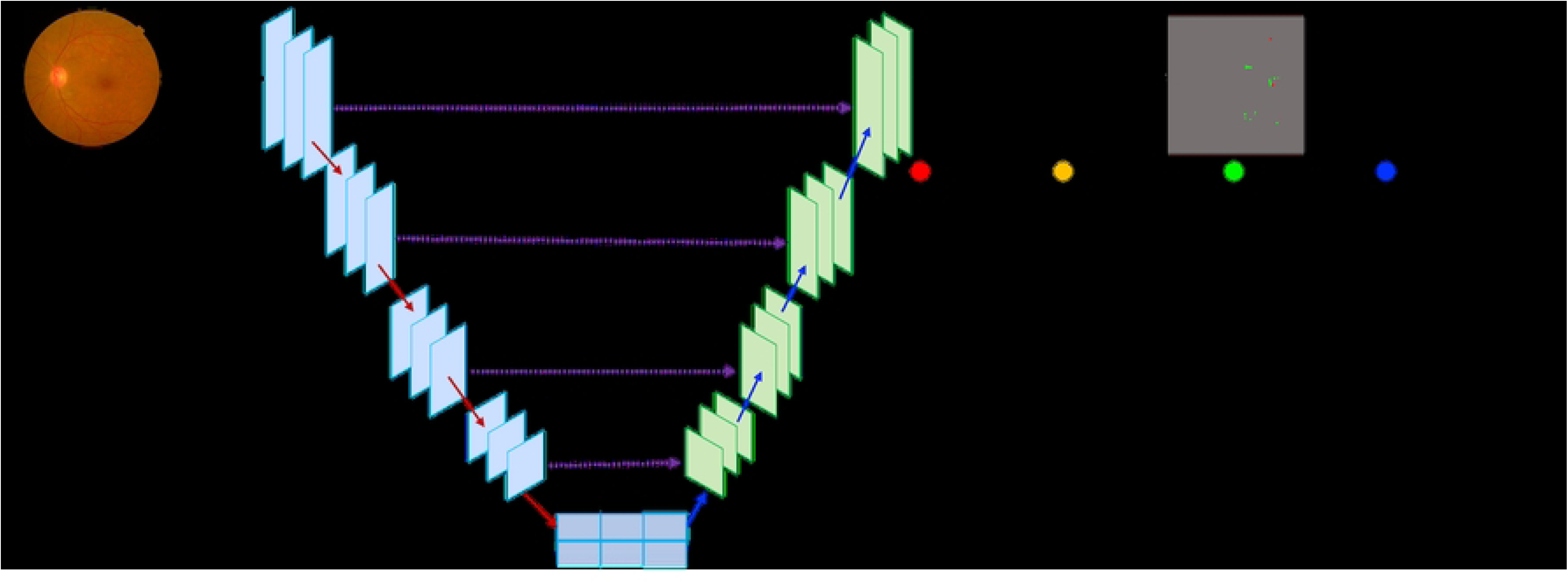
Architecture of the deep learning model for lesion-level detection in fundus images. A U-Net–based convolutional neural network was developed to detect four diabetic retinopathy lesions: hemorrhages, hard exudates, cotton wool spots, and microaneurysms. Each fundus image was processed through a shared encoder–decoder structure with skip connections to produce four lesion-specific segmentation masks. Ground truth annotations were prepared separately for each lesion type. The model was trained using five-fold cross-validation. This architecture enables interpretable, per-lesion outputs that are spatially localized and align with the diagnostic patterns used by ophthalmologists.

**Fig 2.**
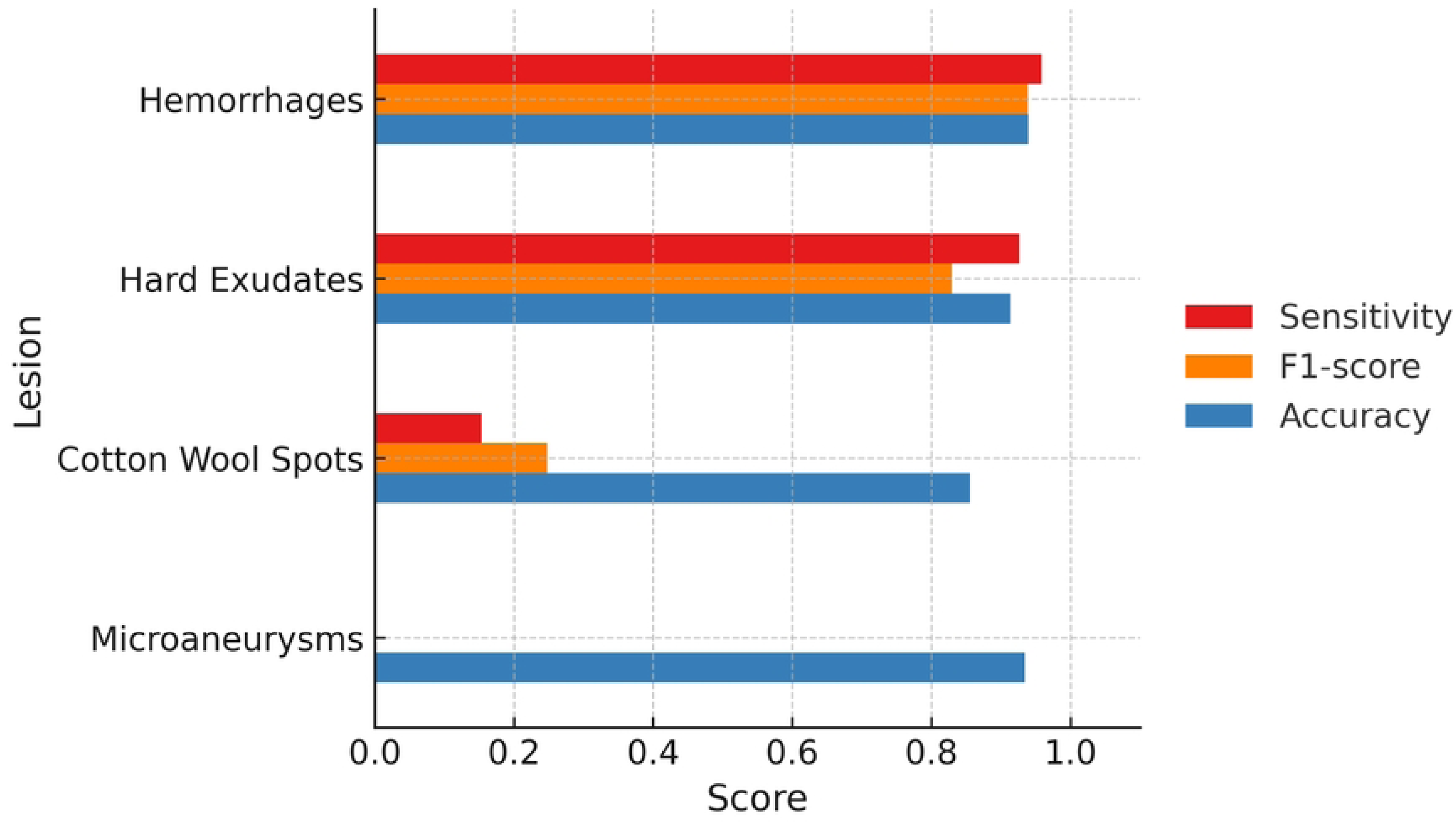
Per-lesion performance metrics of the deep learning model for diabetic retinopathy detection. Bar graph showing the sensitivity, precision, F1 score, and accuracy for detecting four diabetic retinopathy lesions: hemorrhages, hard exudates, cotton wool spots, and microaneurysms. Metrics were derived from a five-fold cross-validated model evaluated on 632 fundus images from patients with mild-to-moderate nonproliferative diabetic retinopathy (DR1 and DR2). The model achieved high performance for hemorrhages and hard exudates, moderate performance for cotton wool spots, and very low sensitivity for microaneurysms. Despite this, the accuracy for detecting microaneurysms remained relatively high due to class imbalance and a large number of true negatives.

**Fig 3.**
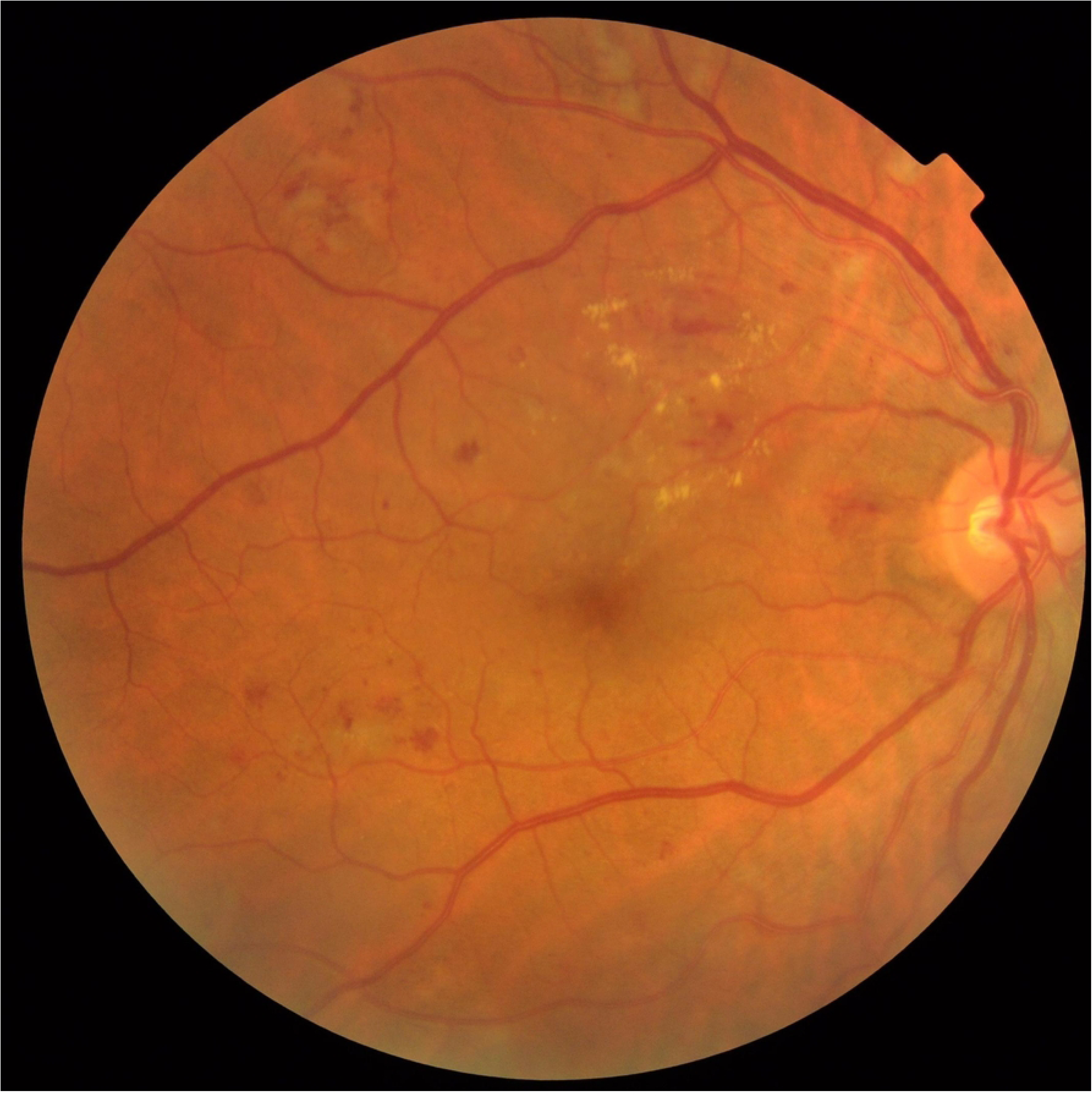
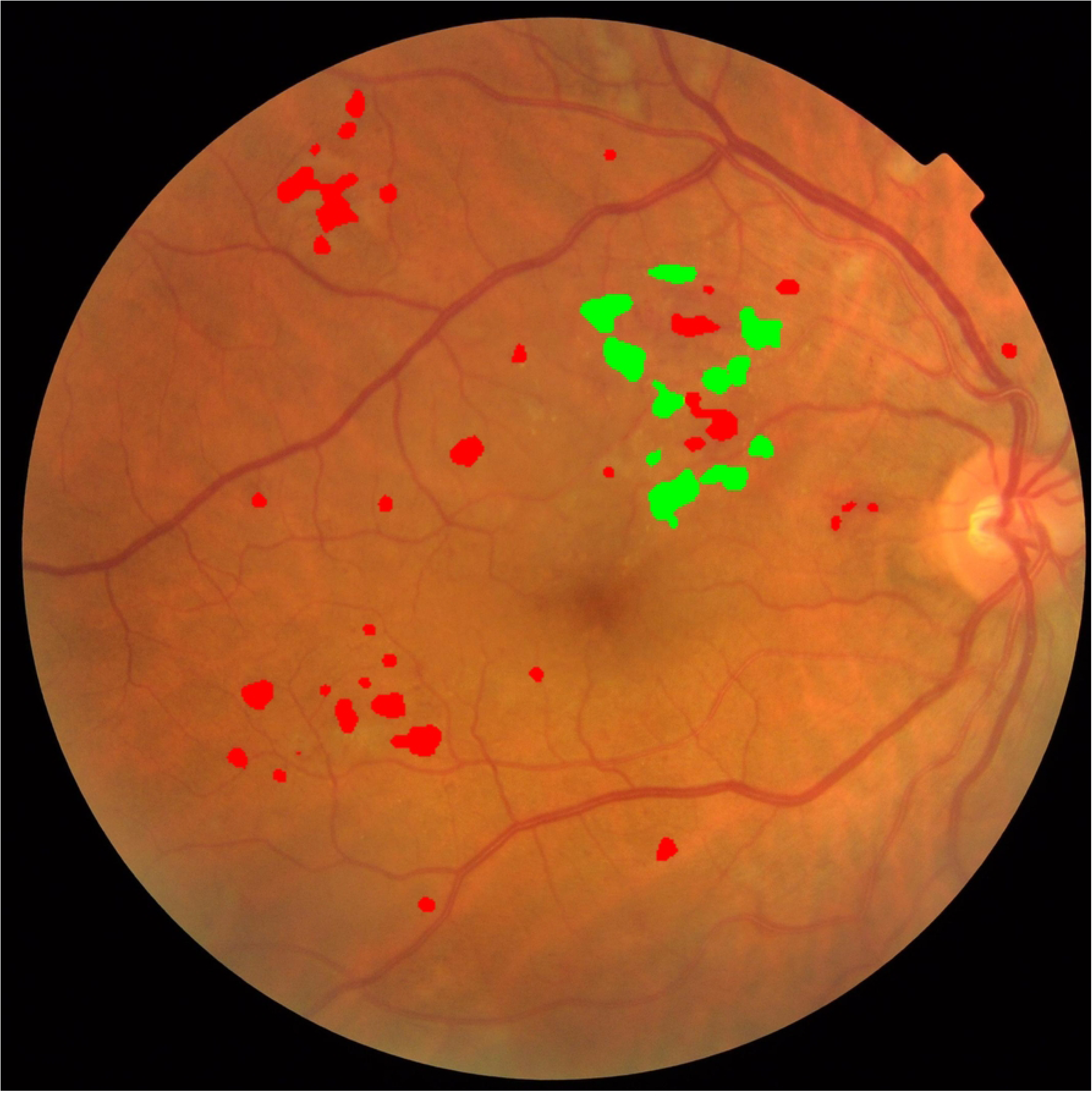
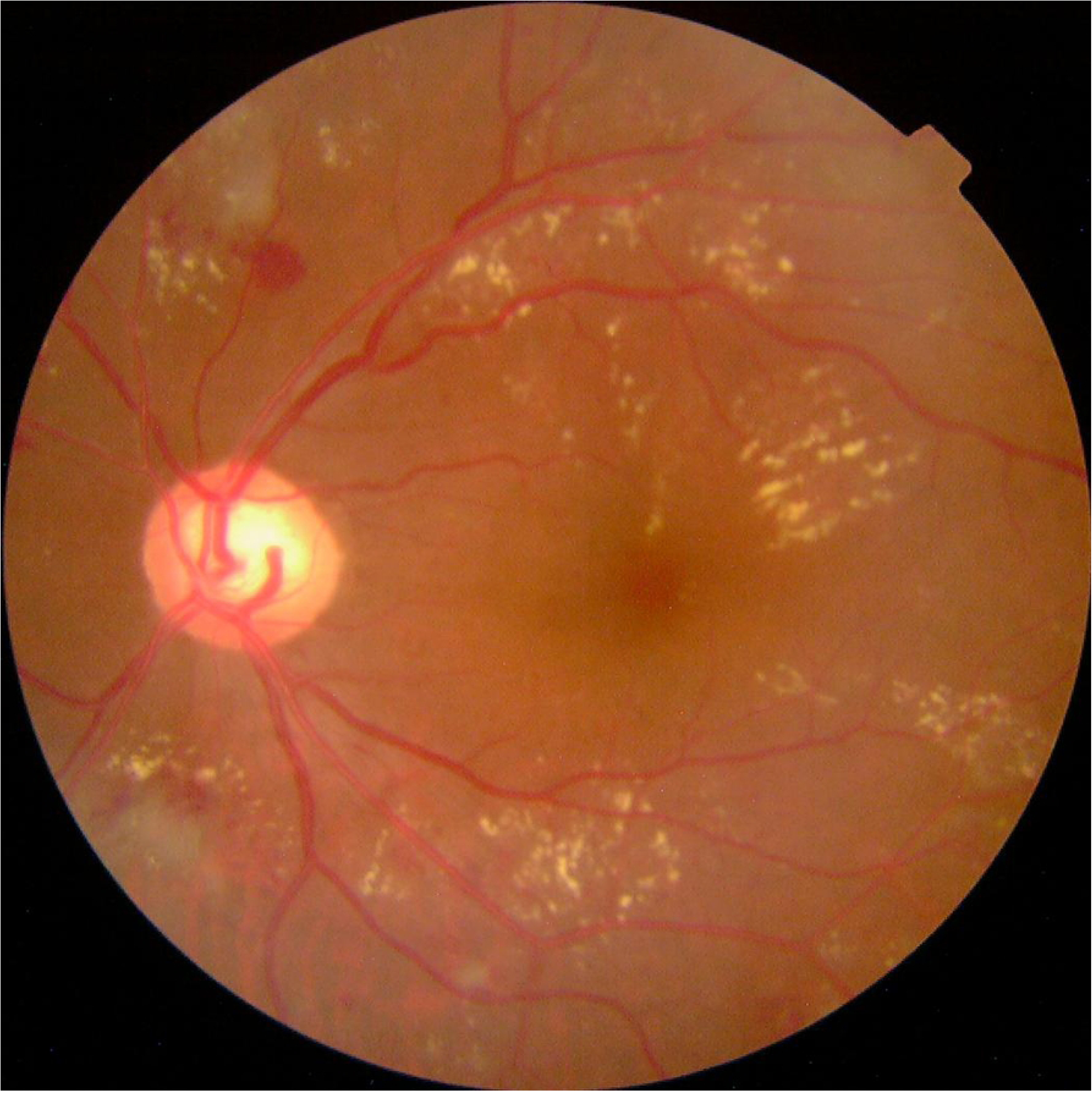
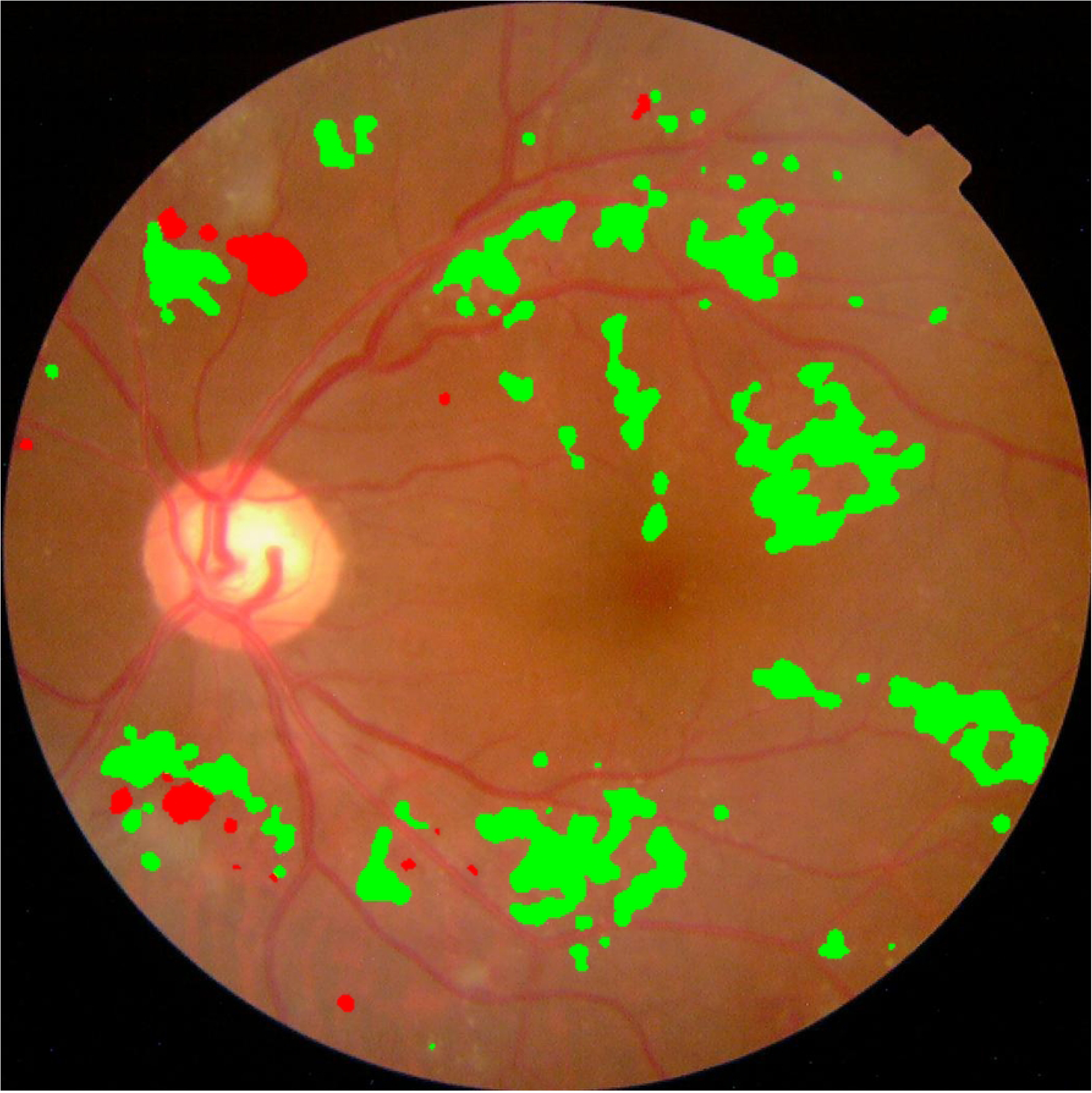
Representative examples of successful lesion detection using an artificial intelligence (AI) model. Two representative cases demonstrating successful lesion-level detection by an AI model. (A) A fundus image graded as DR2 (moderate nonproliferative diabetic retinopathy) containing both hemorrhages and hard exudates. (B) The corresponding AI-generated prediction map showing accurate detection of both lesion types. (C–D) A second DR2 case exhibiting both hemorrhages and hard exudates, with full agreement between the model prediction and the ground truth. All lesions were correctly detected in both cases, resulting in strict matches between AI output and ground truth annotations.

**Fig 4.**
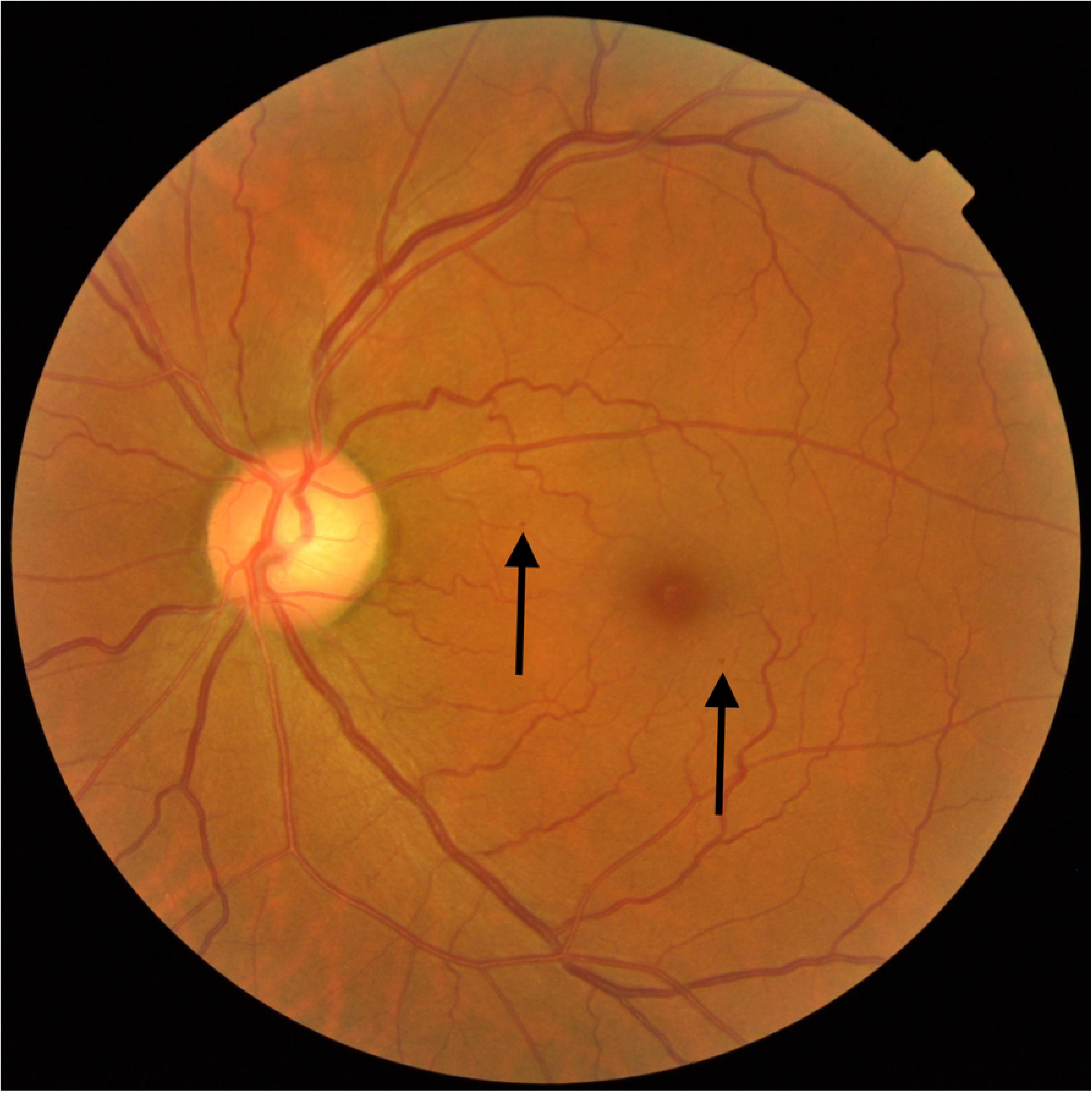
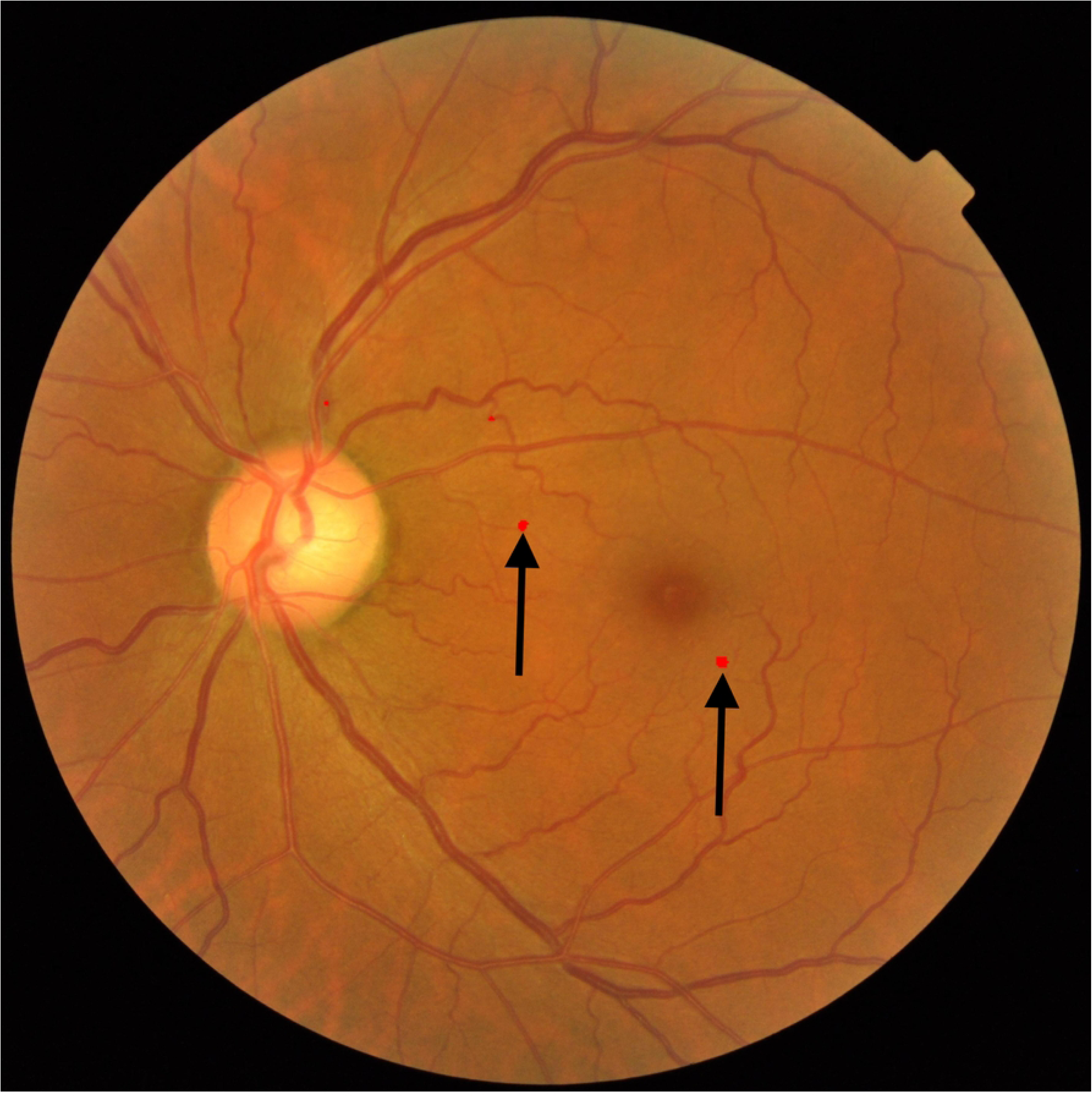
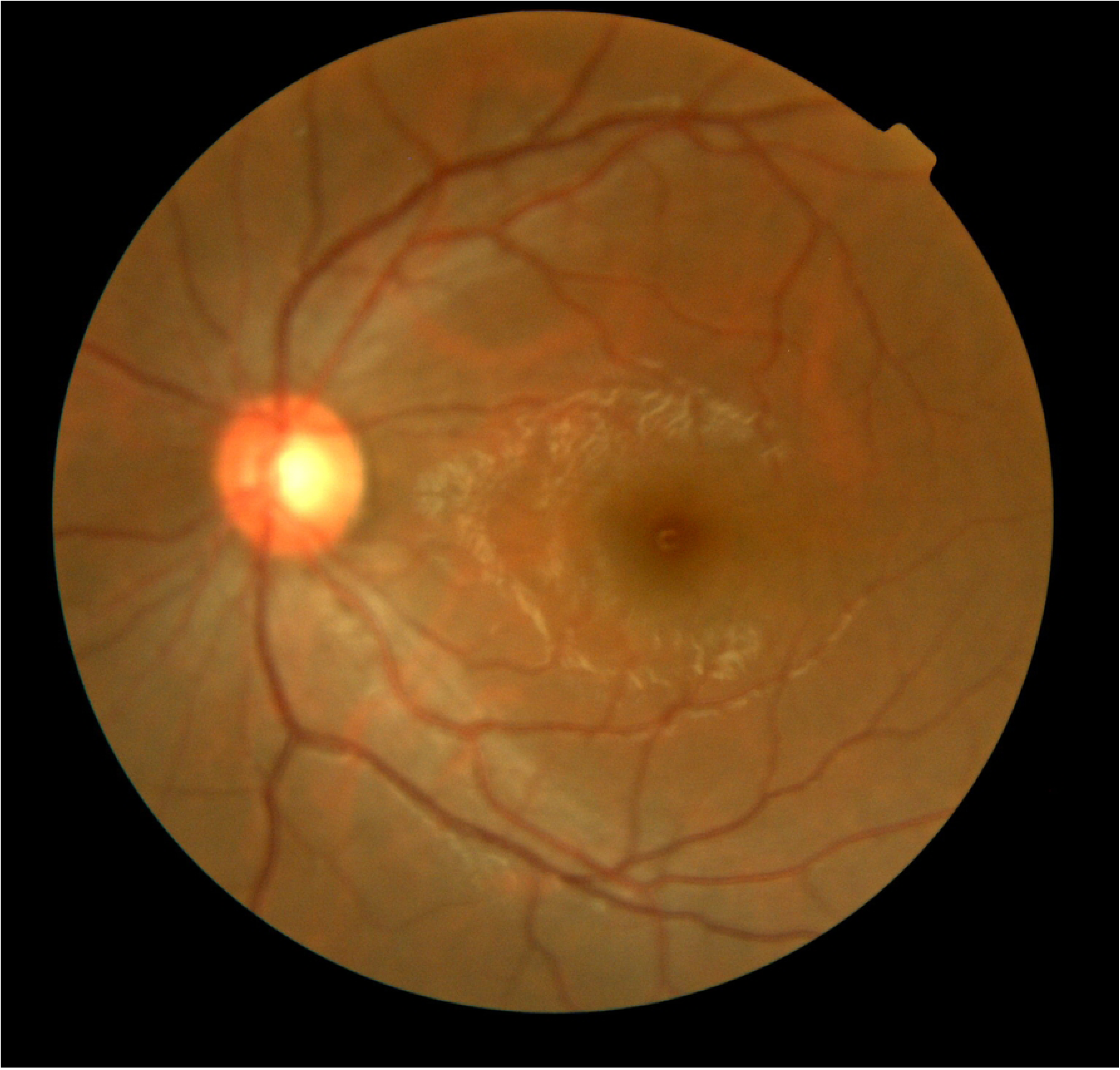
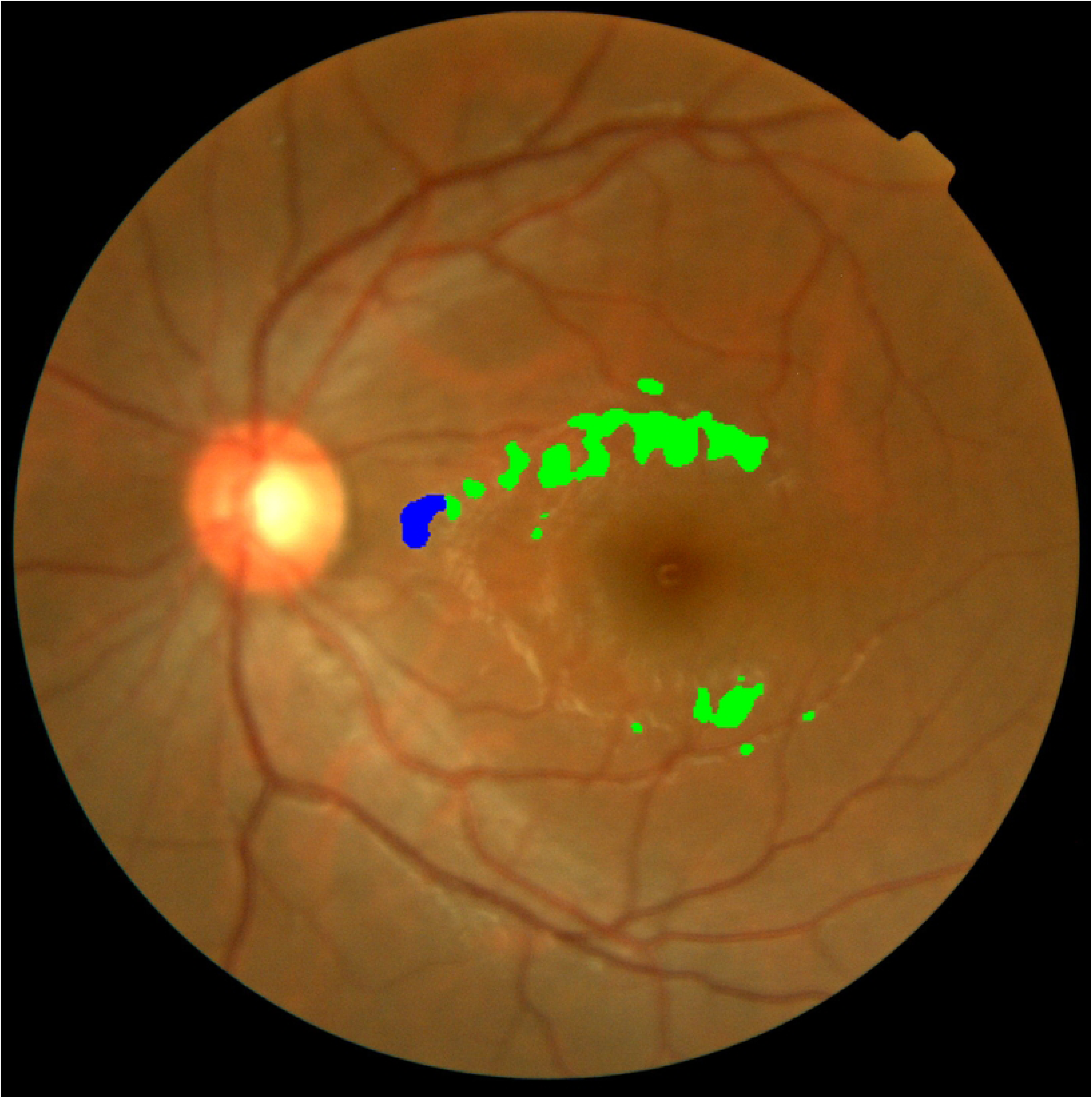
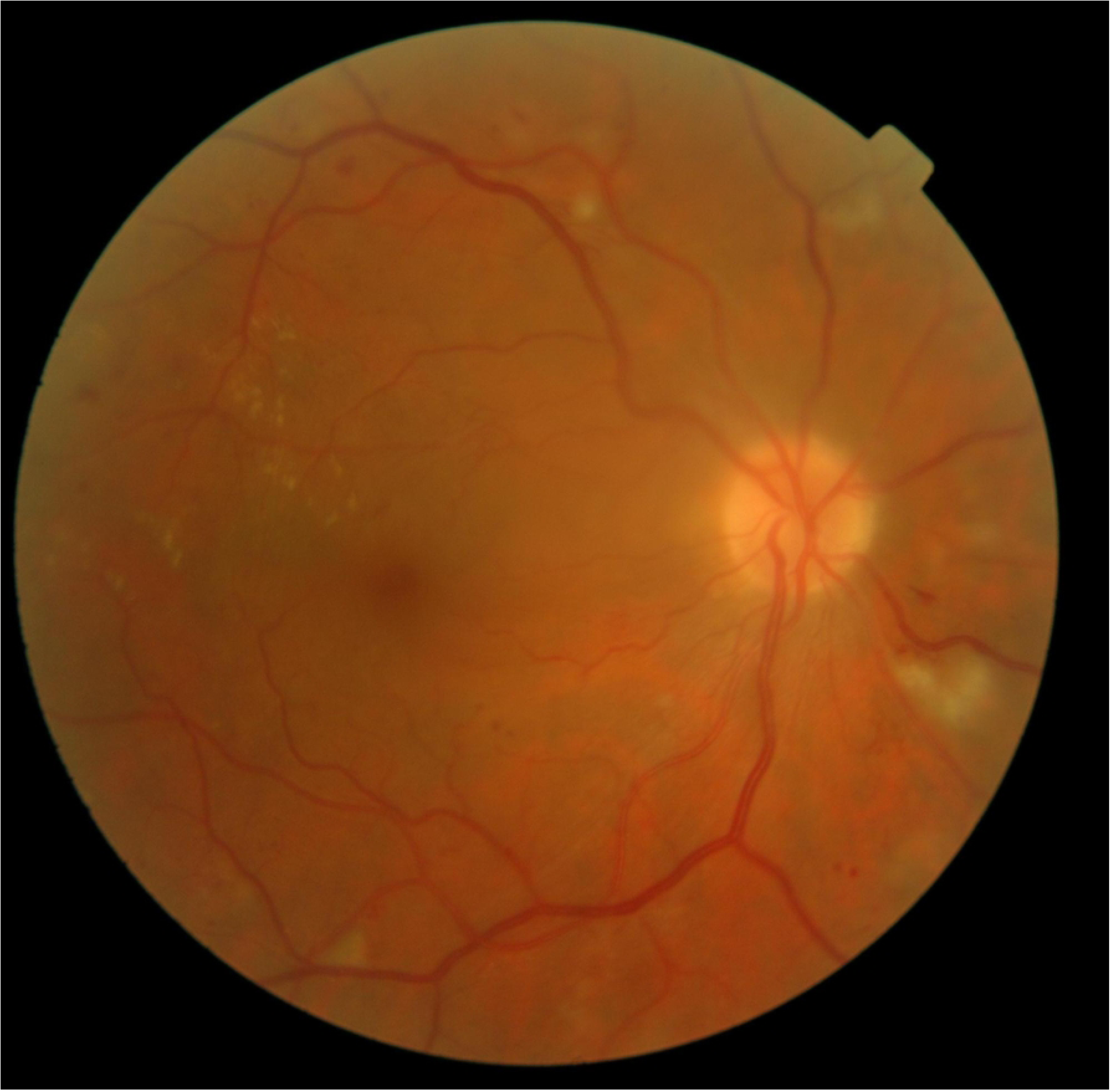
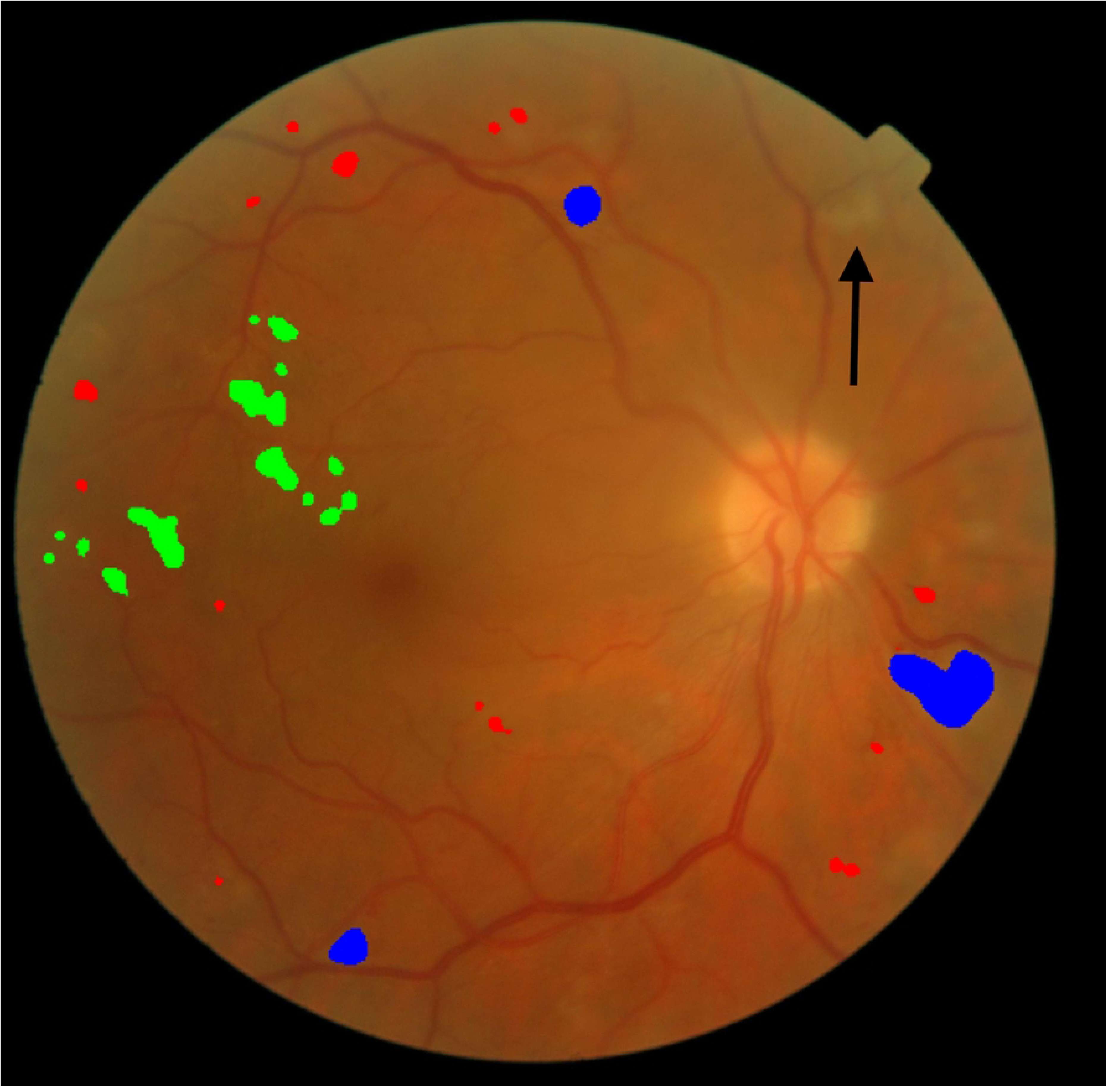
Representative examples of lesion misclassification using an artificial intelligence (AI) model. Three representative cases illustrating typical patterns of lesion misclassification and partial detection errors in the AI model. (A) A fundus image graded as DR1 (mild nonproliferative diabetic retinopathy) containing a microaneurysm, with an arrow indicating the lesion. (B) The corresponding AI prediction misclassified the microaneurysm as a hemorrhage and falsely identified two vessel reflexes as hemorrhages. This case represents an “any-lesion match,” in which the model failed to identify the correct lesion type but still recognized the abnormal image. (C) A DR0 fundus image (no diabetic retinopathy) exhibiting a physiological ring-shaped reflex with no annotated lesions. (D) The AI misinterpreted the reflexes as hard exudates or cotton wool spots, resulting in false positive predictions. This case was evaluated as a complete mismatch across all syntactic agreement criteria. (E) A fundus image graded as DR2 (moderate nonproliferative diabetic retinopathy ) with multiple annotated lesions: hemorrhages, hard exudates, and cotton wool spots. (F) The AI prediction correctly detected all lesion types but partially misclassified a cotton wool spot lesion (indicated by an arrow). Although this case achieved a strict match in terms of lesion categories, it highlights the model’s limitations in spatial completeness.

**Table 2.** Lesion-wise detection performance of the deep learning model for diabetic retinopathy.

| Lesion | Sensitivity | Specificity | Precision | F1 score | Accuracy |
| --- | --- | --- | --- | --- | --- |
| Hemorrhages | 0.9497 | 0.9358 | 0.9327 | 0.9406 | 0.9420 |
| Hard exudates | 0.9253 | 0.9102 | 0.7458 | 0.8259 | 0.9135 |
| Cotton wool<br>spots | 0.1529 | 0.9836 | 0.6341 | 0.2464 | 0.8537 |
| Microaneurysms | 0.0000 | 1.0000 | - | 0.0000 | 0.9356 |

Summary of lesion-wise detection performance metrics for the deep learning model, including sensitivity, specificity, precision, F1-score, and accuracy. Results are reported for four types of diabetic retinopathy lesions: hemorrhages, hard exudates, cotton wool spots, and microaneurysms. Metrics were calculated using a five-fold cross-validated model evaluated on 632 fundus images from patients with mild-to-moderate nonproliferative diabetic retinopathy (DR1 and DR2). The model achieved high performance for hemorrhages and hard exudates, whereas the detection of cotton wool spots was less reliable. Microaneurysms were not correctly detected in any of the cases, resulting in a sensitivity and F1 score of 0.0. Precision is not defined in the absence of true positive predictions, and it is indicated as “—”. The apparent high accuracy of microaneurysms reflects the high number of true negatives.

### Syntactic-Level Agreement Analysis

To further evaluate the model’s clinical utility, we performed a syntactic-level agreement analysis by comparing the predicted and ground truth lesion sets on a per-image basis. This approach simulates how clinicians interpret fundus photographs—not by reproducing each lesion exactly but by identifying key pathological patterns that inform diagnosis. Five syntactic-level agreement metrics were applied: strict match, syntactic containment, hemorrhage match, lesion match, and any-lesion match. These criteria reflect varying degrees of leniency and aim to capture clinically relevant agreement even when the lesion classification is imperfect. Table 3 summarizes the match rates across different diabetic retinopathy severity levels (DR0, DR1, and DR2). The results revealed that although strict match rates declined with increasing lesion complexity, more permissive criteria such as hemorrhage match and any-lesion match rates remained consistently high. To facilitate interpretation, Supplementary Figure S2 presents full 16 × 16 binary matrices of the syntactic outcomes across all lesion-set combinations, visually illustrating the overlap and relative strictness of each metric.

**Table 3.**
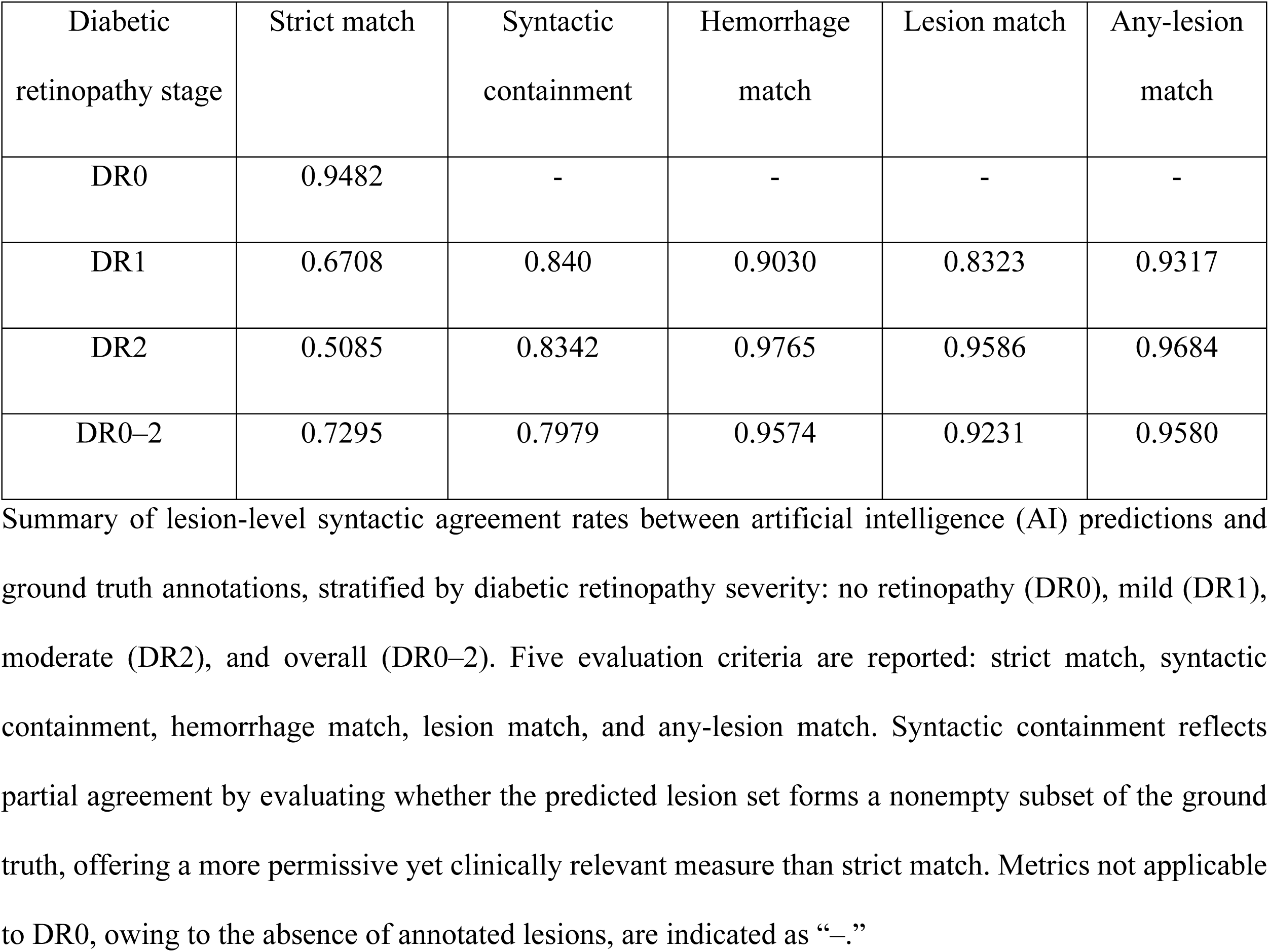
Syntactic match rates according to diabetic retinopathy stage, evaluated based on five agreement criteria. Metrics for images without retinopathy were partially omitted due to the absence of lesions.

### Qualitative Analysis of Misclassifications

Among the 70 ground truth-positive cases of microaneurysms, none were correctly identified by the model. Notably, 70 of these cases (92.9%) were misclassified as hemorrhages, suggesting that although the model responded to the visual presence of microaneurysms, it failed to distinguish them as a separate lesion category. This finding highlights a critical weakness in lesion-specific discrimination. Similar confusion was observed for other lesion types. Cotton wool spots were frequently misclassified as hard exudates, which could be attributed to their similar brightness and texture profiles. Additionally, physiological retinal reflexes—especially in younger individuals—were sometimes misclassified as cotton wool spots or hard exudates. As shown in Figure 4, common misclassification patterns included the following: (A–B) microaneurysms misclassified as hemorrhages, (C–D) cotton wool spots misclassified as hard exudates, (E–F) a physiological reflex in a DR0 case identified as soft exudate, and (G–H) a vessel reflex misinterpreted as hard exudates. These examples illustrate the model’s difficulty in distinguishing subtle or visually similar features.

## Discussion

In this study, we developed and evaluated a deep learning model for detecting four major diabetic retinopathy-related lesions—hemorrhages, hard exudates, cotton wool spots, and microaneurysms—from fundus images. To evaluate the performance of the proposed model, we used lesion-wise metrics and a novel syntactic agreement framework that evaluates lesion-set consistency at the image level. This dual evaluation strategy aimed to capture both granular detection performance and clinically meaningful agreement with expert interpretation.

The syntactic agreement framework offered complementary perspectives for interpreting the model’s clinical utility. For example, although the model showed zero sensitivity for microaneurysms, it responded to most microaneurysms-positive cases by misclassifying them as hemorrhages. Consequently, the any-lesion match rate exceeded 95%, indicating that the model often flagged subtle abnormalities, even when the lesion classification was inaccurate. These findings underscore the practical value of deep learning models in diabetic retinopathy screening, where the primary objective is to minimize missed cases that require further medical evaluation. In real-world screening workflows, such as routine ophthalmologic examinations, the model’s ability to detect lesions indicative of diabetic retinopathy— even when the specific lesion type is not perfectly classified—can effectively support clinicians in identifying patients who may benefit from additional diagnostic testing.

High sensitivity and F1 scores were observed for hemorrhages and hard exudates. This strong performance can be attributed to their relatively distinct visual characteristics (such as shape, size, and contrast) and their higher prevalence within the training dataset. These factors make it easier for deep learning models to learn and recognize, and they are also more consistently annotated by human readers.

In contrast, the detection of cotton wool spots and microaneurysms proved substantially more challenging. Cotton wool spots showed low sensitivity, possibly due to their variable size and diffuse appearance. Microaneurysms were almost entirely missed, likely due to their small size, low contrast, and visual similarity to background artifacts. These characteristics make such lesions difficult to reliably detect for both human experts and AI systems, highlighting a critical limitation in the sensitivity of the current model to subtle lesions.

The visual misclassification patterns observed in this study reflect common challenges in fundus image analysis. Cotton wool spots were often misclassified as hard exudates because of their overlapping texture and brightness, while physiological retinal reflexes—particularly in younger patients—were sometimes misclassified as pathological lesions. These findings highlight the limitations of current segmentation models in distinguishing pathological retinal features from nonpathological ones and highlight the need for more diverse training data and refined model architectures.

Prior pixel-level lesion-aware segmentation models have primarily relied on per-lesion performance metrics, such as sensitivity, specificity, and F1 score [6,7]. Although these metrics are informative, they do not fully capture the holistic diagnostic reasoning used in clinical practice, where multiple lesion types are integrated in a comprehensive assessment. Although some recent studies have proposed attention-based or region-focused methods to approximate this process [8,9], none of them have introduced a quantitative framework to evaluate the internal consistency of lesion predictions at the image level. Our syntactic agreement framework addresses this gap by introducing structured, interpretable metrics that are more closely aligned with how clinicians assess fundus photographs in real-world settings.

This study had several limitations. First, lesion annotations were performed by a single annotator under ophthalmologist supervision. Although this approach ensured consistency, it may have introduced labeling bias, especially for subtle or borderline findings, such as microaneurysms or cotton wool spots. Second, the model exhibited poor sensitivity for microaneurysms, likely due to a combination of factors, including class imbalance, limited visual distinctiveness, and architectural constraints. Third, although syntactic agreement metrics are effective in approximating clinical impressions, they do not penalize incorrect lesion types if any abnormality is detected. This may lead to an overestimation of model reliability, particularly in applications requiring precise lesion classification.

Despite these limitations, our findings have important clinical implications. By demonstrating that deep learning-based segmentation can achieve high syntactic agreement with expert-level lesion patterns—even when the exact classification is imperfect—this study highlights the potential of syntactic evaluation to bridge the gap between AI predictions and clinical expectations. Future research should focus on validating this framework across larger, multi-institutional datasets, exploring its integration into real-world screening workflows, and enhancing model architectures to improve the detection of subtle lesion types.

## Conclusion

We developed an explainable deep learning model for the lesion-level detection of diabetic retinopathy using fundus images and proposed a novel syntactic agreement framework to evaluate prediction consistency at the image level. Although the lesion-level metrics revealed high detection performance for hemorrhages and hard exudates, the model showed moderate performance for cotton wool spots and failed to detect microaneurysms reliably, with the latter being frequently misclassified. However, syntactic agreement metrics such as any-lesion match and hemorrhage match rates remained consistently high, indicating that the model can identify abnormal images even when the lesion-type classification was imperfect. By integrating lesion-wise and syntactic-level evaluations, this study provides a more comprehensive and clinically relevant assessment of AI performance in retinal image analysis. The proposed syntactic agreement framework provides a practical tool for evaluating the clinical reliability of AI outputs in diabetic retinopathy screening scenarios, where the detection of key abnormalities is often more important than perfect lesion classification. Future work should focus on validating this framework across diverse clinical settings and further enhancing model sensitivity to subtle lesion types to ensure safe and effective deployment in real-world environments.

## Data Availability

This study used publicly available datasets from Kaggle competitions: the APTOS 2019 Blindness Detection dataset (https://www.kaggle.com/competitions/aptos2019-blindness-detection) and the Diabetic Retinopathy Detection dataset provided by EyePACS (https://www.kaggle.com/competitions/diabetic-retinopathy-detection). Both datasets consist of de-identified fundus images. No additional data were generated or analyzed during the current study.

https://www.kaggle.com/competitions/aptos2019-blindness-detection

## Acknowledgments

We thank EyePACS and the organizers of the APTOS 2019 Blindness Detection Challenge for providing the publicly available dataset used in this study.

---

**Supplementary Fig S1. Manual annotation interface and labeling workflow.**

This figure illustrates the annotation interface and manual labeling process used to generate lesion-level ground truth in fundus images. The left panel shows the initial fundus image with highlighted lesion candidates. On the right, multiple labeling operations are demonstrated: lesion selection by mouse click, region outlining for grouped annotation, and line-based editing to generate or erase lesion masks. Each lesion type was individually annotated to support lesion-wise model training and evaluation.

**Supplementary Fig S2. Binary matrices of syntactic agreement outcomes across all combinations of diabetic retinopathy lesion sets.**

Each panel (A–E) shows a 16×16 binary matrix comparing combinations of lesion types between ground truth and AI output. Lesions include hemorrhages (H), hard exudates (HE), cotton wool spots (CWS), and microaneurysms (MA). For each syntactic agreement criterion—Strict Match (A), Syntactic Containment (B), Lesion Match (C), Any-Lesion Match (D), and Hemorrhage Match (E)—cells are color-coded to indicate whether the criterion was met (red), not met (blue), or not applicable (blank). These matrices enable visual comparison of how stringently each criterion evaluates lesion-wise agreement between ground truth and AI output.

Abbreviations:

H, hemorrhages; HE, hard exudates; CWS, cotton wool spots; MA, microaneurysms; AI, artificial intelligence; DR, diabetic retinopathy.

**Supplemental Table S1. Lesion-wise detection performance in DR1**

Detection performance metrics for each lesion type on DR1 fundus images. Metrics include sensitivity, specificity, precision, F1-score, and accuracy. No true positives were observed for microaneurysms, resulting in a precision value that is undefined.

**Supplemental Table S2. Lesion-wise detection performance in DR2.**

Detection performance metrics for each lesion type on DR2 fundus images. Metrics include sensitivity, specificity, precision, F1-score, and accuracy. No true positives were observed for microaneurysms, resulting in a precision value that is undefined.

## Notes

### Competing Interest Statement

The authors declare no competing interests.

### Funding Statement

The author(s) received no specific funding for this work.

### Author Declarations

IRB or oversight body name: Not applicable – study used only publicly available, de-identified datasets IRB approval number or exemption statement: This study used only publicly available, de-identified data from the APTOS and EyePACS datasets hosted on Kaggle. Therefore, ethical approval and patient consent were not required in accordance with institutional and journal policies.

